# Short term heart rate variability is preserved in Parkinson’s disease under atomoxetine

**DOI:** 10.64898/2026.03.15.26348415

**Authors:** Isabella F. Orlando, Frank H. Hezemans, Kamen A Tsvetanov, Rong Ye, Catarina Rua, Ralf Regenthal, Roger A. Barker, Caroline H. Williams-Gray, Luca Passamonti, Trevor W. Robbins, James B. Rowe, Claire O’Callaghan

## Abstract

Repurposed noradrenergic drugs have been proposed to treat neuropsychiatric symptoms in Parkinson’s disease and related conditions. While there is evidence that these drugs can be beneficial for cognition in selected patients, questions remain about their cardiovascular effects. Here we tested whether heart rate variability (HRV) is altered in people with Parkinson’s disease, following a single-dose challenge with the noradrenaline reuptake inhibitor atomoxetine (40 mg, oral). Consistent with previous work, our cohort of people with idiopathic Parkinson’s disease (n=15) had lower HRV than healthy controls (n=22). Decreased HRV in people with Parkinson’s disease was associated with reduced integrity of the caudal locus coeruleus, measured using neuromelanin-sensitive ultra-high field 7T magnetic resonance imaging. Following a randomised double-blind placebo-controlled crossover challenge in the Parkinson’s disease group, short-term resting HRV was not significantly altered following atomoxetine. Using Bayesian statistical inference, we demonstrated confidence in the preservation of HRV across measures in the time, frequency, and non-linear domains. Our findings are in favour of a safe cardiovascular profile for atomoxetine in Parkinson’s disease, further supporting noradrenergic modulation as a viable treatment strategy for neuropsychiatric symptoms in Parkinson’s disease and related disorders.

## Introduction

The noradrenergic locus coeruleus is one of the earliest sites of pathology in Parkinson’s disease and is implicated in both motor and non-motor symptoms.^1–3^ Repurposed noradrenergic drugs have been reported to improve cognitive and psychiatric symptoms in some individuals,^4–8^ with the possibility that they may also have disease-modifying effects.^9,10^ However, autonomic problems in Parkinson’s disease raise critical questions about the safety and cardiovascular effects of noradrenergic treatment approaches.

Of particular interest is the noradrenaline reuptake inhibitor atomoxetine, which increases extracellular levels of noradrenaline^11^ and phasic-to-tonic locus coeruleus firing.^12^ Atomoxetine is licensed in many countries for the treatment of attention deficit hyperactivity disorder in children and adults. The cardiovascular safety of atomoxetine is well described in these patient groups. The drug is generally very well-tolerated and associated with only small increments in blood pressure or pulse rate^13^ that are not linked with serious adverse cardiac events.^14,15^ Atomoxetine is tolerated well in Parkinson’s disease following acute challenges^5,7,8^ and longer-term trials.^16,17^ However, potential cardiovascular changes under atomoxetine in Parkinson’s disease are not well documented. This is particularly relevant given the cardiac and autonomic symptoms associated with Parkinson’s disease.

Noradrenergic sympathetic innervation of the heart is affected in Parkinson’s disease, resulting in profound myocardial noradrenaline deficiency.^18^ Cardiac sympathetic denervation (e.g., measured by ^123^I-metaiodobenzylguanidine, MIBG scintigraphy) supports the diagnosis of Parkinson’s disease^19^ and progresses with disease severity.^20^ Nuclei that provide central control of cardiac function are affected in Parkinson’s disease, including the locus coeruleus, which plays a major role in cardiovascular regulation.^21^ Cardiovascular autonomic dysfunction in Parkinson’s disease can manifest as abnormal blood pressure (e.g., labile blood pressure or orthostatic hypotension),^22^ or changes in the dynamic, adaptive regulation of heart rhythms. The latter is captured by measures of heart rate variability (HRV), which is reduced in Parkinson’s disease.^23^

Here, we measured short-term resting HRV in people with Parkinson’s disease undergoing an acute, single-dose challenge with 40 mg oral atomoxetine or placebo. Given the tolerability of atomoxetine in Parkinson’s disease, and its mild cardiovascular profile in attention deficit hyperactivity disorder, we predicted that we would not see significant HRV changes under the drug. Bayesian statistical inference was used given the predicted null effect and given its advantages in a small sample size, including the incorporation of priors and generation of full posterior distributions that allow for a more direct quantification of uncertainty. Analyses first focussed on the atomoxetine vs. placebo comparison (i.e., the drug effect), and then on comparisons between Parkinson’s disease (on placebo) vs. healthy controls (i.e., interpreted as a disease effect). Participants underwent neuromelanin-sensitive neuroimaging with 7T MRI, providing a marker of locus coeruleus integrity. We compared locus coeruleus integrity against the HRV metrics that were altered in people with Parkinson’s compared to controls, to determine whether disease-related changes in HRV relate to reduced locus coeruleus integrity.

## Materials and Methods

General study design and participant characteristics overlap with previous publications.^4,6,7,24^ The study was approved by the local Ethics Committee (REC 10/H0308/34) and participants provided written informed consent according to the Declaration of Helsinki. The study is registered on the ISRCTN registry with study ID ISRCTN46299660 (https://doi.org/10.1186/ISRCTN46299660).

### Participants

Nineteen individuals with Parkinson’s disease were recruited through the University of Cambridge Parkinson’s disease research clinic and Parkinson’s UK volunteer panels. Participants met United Kingdom Parkinson’s Disease Society Brain Bank criteria, were aged between 50–80 years, with Hoehn and Yahr stages 1.5–3, no dementia and no contraindications to atomoxetine or 7T MRI. Twenty-six age-, sex- and education-matched healthy control participants were recruited from local volunteer panels. Control participants were screened for a history of psychiatric or neurological disorders and were not taking psychoactive or noradrenergic medications. Following ECG data preprocessing and signal-quality checks (described below), the final sample analysed included 15 people with Parkinson’s disease and 22 controls.

### Study procedure

Participants with Parkinson’s disease were tested over three sessions. Session one included MRI scanning and baseline clinical assessment of cognition and motor function (see Table 2). Sessions two and three were a double-blind, placebo-controlled crossover design, with participants randomised to receive 40mg of atomoxetine or placebo. The visits were scheduled ≥6 days apart [mean = 7.4 days; standard deviation (SD) 1.8 days; range 6-14 days]. At each visit, blood samples were taken 2 hours post drug/placebo administration, corresponding to the predicted peak in plasma concentration following an oral atomoxetine dose.^25^ Mean plasma concentration^26^ was 264.07 ng/mL after atomoxetine (SD = 124.50 ng/mL, range: 90.92-595.11 ng/mL) and 0 ng/mL after placebo. Participants then commenced a battery of computerised cognitive tasks that took approximately 2 hours; ECG recording was performed during a rest break midway through the testing session. Supine/lying and sitting upright blood pressure and pulse rate measures were monitored three times across the session (on arrival, 2 hours post tablet administration, and on completion of testing). Subjective measures of mood were collected prior to tablet administration and 2 hours post, using a set of 16 visual analogue scales.^27^ All sessions were completed at a similar time of day, with participants on their regular anti-parkinsonian medications. Control participants provided normative data and did not undergo the atomoxetine/placebo manipulation. They were tested in a single session that included MRI scanning and the same experimental task battery undertaken by the Parkinson’s disease group.

### ECG recording

Ten-minute ECG recordings were collected with participants sitting upright in a chair. ECG was recorded using a BIOPAC MP36 system (BIOPAC Systems, Inc., USA) at a sampling frequency of 1000 Hz. Recordings were obtained using disposable electrodes attached to one forearm and both ankles. Participants were asked to sit still and refrain from talking.

### ECG data preprocessing and extraction of heart rate variability metrics

ECG data preprocessing and extraction of HRV metrics used the PhysioNet Cardiovascular Signal Toolbox,^28^ implemented in MATLAB (R2019b). The toolbox prepares ECG recordings for HRV estimation via detection and removal of data based on arrhythmias (including automatised detection of atrial fibrillation and premature ventricular contraction), noise and low signal quality. Data was analysed in a single 10-minute window. Using the default parameter settings as a guide^28^ we set less conservative thresholds to account for possible signal degradation in an older, neurodegenerative disease cohort (see Supplementary Material for parameter settings). Based on these criteria, four healthy controls and four participants with Parkinson’s disease (for both their placebo and atomoxetine sessions) were excluded. Heart rate variability metrics were then calculated on the normal-to-normal (NN) intervals, including time-domain, frequency-domain and non-linear metrics. Time-domain metrics quantify the amount of variability in the inter-beat intervals; frequency-domain metrics estimate absolute or relative power within frequency bands; non-linear metrics quantify the unpredictability, or complexity, of the time series.^29^ See Table 1 and Supplementary Material for descriptions of each metric.

**Table 1.**
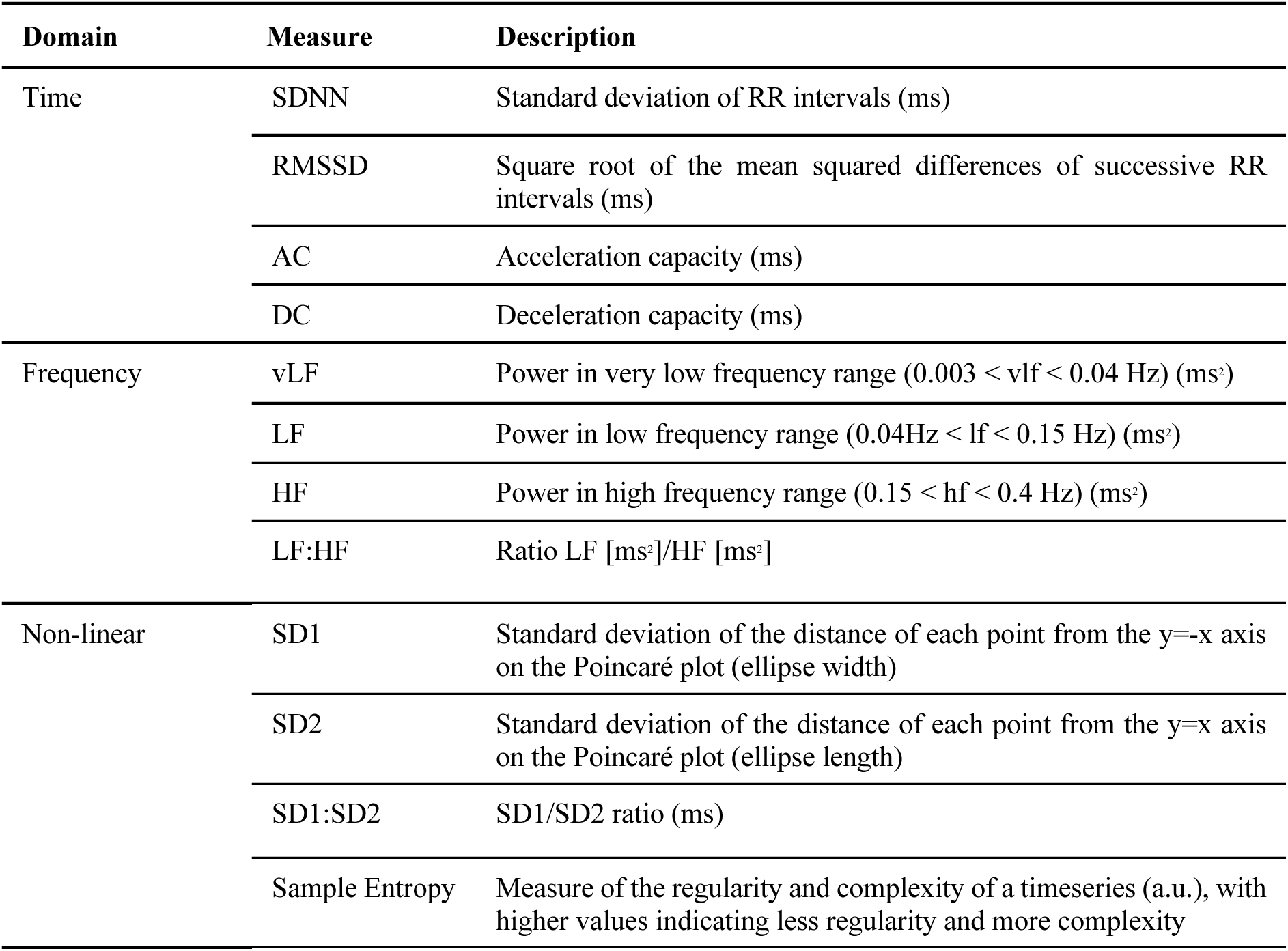
Heart rate variability metrics.

### Statistical analysis

We performed Bayesian estimation of linear mixed effect models to evaluate group differences in HRV metrics. This approach offers several advantages over frequentist methods that are particularly relevant given our study design and relatively small sample sizes. Specifically, Bayesian inference provides a principled framework for quantifying uncertainty and allows for the incorporation of domain expertise or regularisation via prior distributions. In our case, priors were specified to be “weakly informative”, such that they provide moderate regularisation and help stabilise computation, without imposing strong prior beliefs.

For each HRV metric, we performed two separate analyses. First, to assess drug effects, we compared people with Parkinson’s disease on atomoxetine vs. placebo. Second, to assess disease-related effects, we compared people with Parkinson’s on placebo vs. controls (noting that groups differed by disease, and repeated-measures testing, and the expectation of a possible drug treatment effect). Each model included a fixed effect of condition, operationalised as drug status (atomoxetine vs. placebo) or group (Parkinson’s disease vs. control), depending on the contrast of interest. A random intercept for participants was included to account for repeated measures within individuals. In models testing drug effects, we included a fixed effect of visit order (placebo-first vs. atomoxetine-first) as a covariate of no interest.

Following recommendations for regularising inference in small samples,^30^ we used Student’s t-distributions with three degrees of freedom as priors on all fixed-effect regression coefficients and on the residual standard deviation. For each model, the parameters of the Student’s t priors (location and scale) were adjusted based on the empirical distribution of the outcome variable in the reference group of the comparison. This approach was informed by the default method of prior elicitation used in the *rstanarm* package.^31^ Thus, for models analysing a drug effect, the priors were based on the patient group on placebo; for models analysing a disease-related effect, the priors were based on the control group. For all regression coefficients and for the residual SD, the scale parameter of the prior was set to the empirical SD of the reference group. For the intercept, the prior’s location parameter was set to the empirical mean of the reference group; for all other coefficients, the prior’s location parameter was set to zero.

Model fitting was implemented in R (version 4.2.2) using the *brms* package,^32^ which provides an interface to *Stan*^33^ for Hamiltonian Monte Carlo sampling. We used four chains of 5000 iterations each, discarding the first 1000 iterations as warm-up. Convergence diagnostics, including the potential scale reduction factor (*R̂*), were inspected to ensure validity of the posterior samples. For each regression coefficient, we report the mean of its posterior distribution as the estimated effect size, and the 95% quantile interval as the credible interval (CI), representing a range of plausible effect sizes.

We used the Region of Practical Equivalence (ROPE) to assess evidence of group differences. The ROPE defines a range of values considered negligible – that is, practically equivalent to a null effect.^34^ Here, the ROPE was defined as ± 0.1 times the empirical SD of the control group.^35^ If the posterior mean lies outside the ROPE, there is evidence to accept the hypothesis of a difference between groups; if the CI is outside of the ROPE, the evidence for accepting a difference is strong.^35,36^ If the posterior mean is inside the ROPE, this is evidence in favour of the null (i.e., no difference between groups); if the CI lies entirely within the ROPE, then the null hypothesis of no group difference can be accepted.^35,36^

To compare demographic and clinical measures, in order to confirm patient and control groups were reasonably matched, we conducted Bayesian independent t-tests and contingency table analysis using the *BayesFactor* package.^37^ We report the Bayes factor (BF) for the alternative hypothesis (i.e., non-zero group difference) over the null hypothesis, with BF > 1 indicating relative evidence for the alternative hypothesis and BF > 3 indicating positive evidence for the alternative hypothesis.^38^

### Locus coeruleus integrity estimation and relationship with heart rate variability

As part of this study, participants underwent MRI scanning on a 7 T Magnetom Terra (Siemens, Erlangen, Germany) with a 32-channel head coil (Nova Medical, Wilmington, MA, USA). The locus coeruleus was imaged using a neuromelanin-sensitive 3D magnetisation transfer weighted sequence^39^ at high resolution (0.4 × 0.4 × 0.5 mm^3^). Locus coeruleus integrity was estimated as a contrast-to-noise ratio (CNR), derived using pipelines described in^7,40^ and reported in Supplementary Material.

Previous reports from the current study cohort identified significantly reduced integrity, specifically in the caudal locus coeruleus, in people with Parkinson’s disease vs. controls,^7,41^ which is replicated in other studies using neuromelanin-sensitive MRI.^42,43^ Since the caudal locus coeruleus is implicated in descending vagal projections,^21^ we tested whether caudal locus coeruleus integrity in people with Parkinson’s disease related to HRV measures — focusing on those HRV measures that differed convincingly between Parkinson’s participants and controls. We assessed the correlation using Bayesian tests of linear correlation implemented using the *BayesFactor* package.^37,44^

## Results

### Participant characteristics

Parkinson’s disease and control groups were matched for age, years of education, ACE-R, MMSE and MoCA (all BFs < 1; see Table 2). Within the drug and placebo sessions there was no convincing evidence of a change in pulse rate. There was evidence of slightly elevated blood pressure on atomoxetine for the supine diastolic measure, i.e., placebo [mean (range); SD]: arrival = 70.40 (50 – 84; 9.50); 2-hours = 67.60 (58 – 80; 5.08); completion = 72.67 (59 – 87; 8.00); atomoxetine: arrival = 74.07 (55 – 85; 7.42); 2-hours = 74.33 (58 – 94; 8.50); completion = 78.00 (63 – 98; 10.43). Although this was not considered clinically significant and there was no evidence of elevated blood pressure across lying systolic or sitting measures. There was no change in subjective ratings of mood and arousal levels within the sessions. All pulse rate, blood pressure and subjective rating analyses are described in detail in the Supplementary Material.

**Table 2.** Demographics and clinical assessments of participants in their regular medication state. Data are presented as mean (SD). Comparisons of patient and control groups were performed with Bayesian t-tests. BF = Bayes factor, where >3 indicates substantial evidence of a group difference. MMSE, Mini-Mental State Examination; MoCA, Montreal cognitive assessment; ACE-R Total, revised Addenbrooke’s Cognitive Examination Total score; MDS-UPDRS III, Movement Disorders Society Unified Parkinson’s Disease Rating Scale Part III. Levodopa equivalent daily dose calculated using.^45^

|  | PD (n=15) | Control (n=22) | BF |
| --- | --- | --- | --- |
| Age (years) | 67.20 (7.66) | 65.41 (5.71) | 0.418 |
| Education (years) | 14.27 (1.98) | 14.96 (3.19) | 0.400 |
| Male / Female | 12 / 3 | 12 / 10 | 2.302 |
| MMSE | 29.47 (0.75) | 29.73 (0.55) | 0.578 |
| MoCA | 28.20 (1.78) | 28.68 (1.39) | 0.449 |
| ACE-R Total | 95.53 (3.83) | 97.55 (3.35) | 0.967 |
| MDS-UPDRS III | 27.40 (12.77) |  |  |
| Hoehn and Yahr stage | 2.33 (0.49) |  |  |
| Disease duration (years) | 3.81 (1.55) |  |  |
| Levodopa equivalent daily dose (mg/day) | 521.27 (444.99) |  |  |

### Evidence for preserved heart rate variability in Parkinson’s disease on atomoxetine vs. placebo

HRV metrics for the three groups are shown in Figure 1 (see Supplementary Table 2 for group means). For the Parkinson’s disease atomoxetine vs. placebo comparisons, the posterior distributions of HRV estimates and ROPE are shown in Figure 2. Overall, there was good evidence in favour of the null hypothesis of no difference between the atomoxetine vs. placebo sessions.

**Figure 1.**
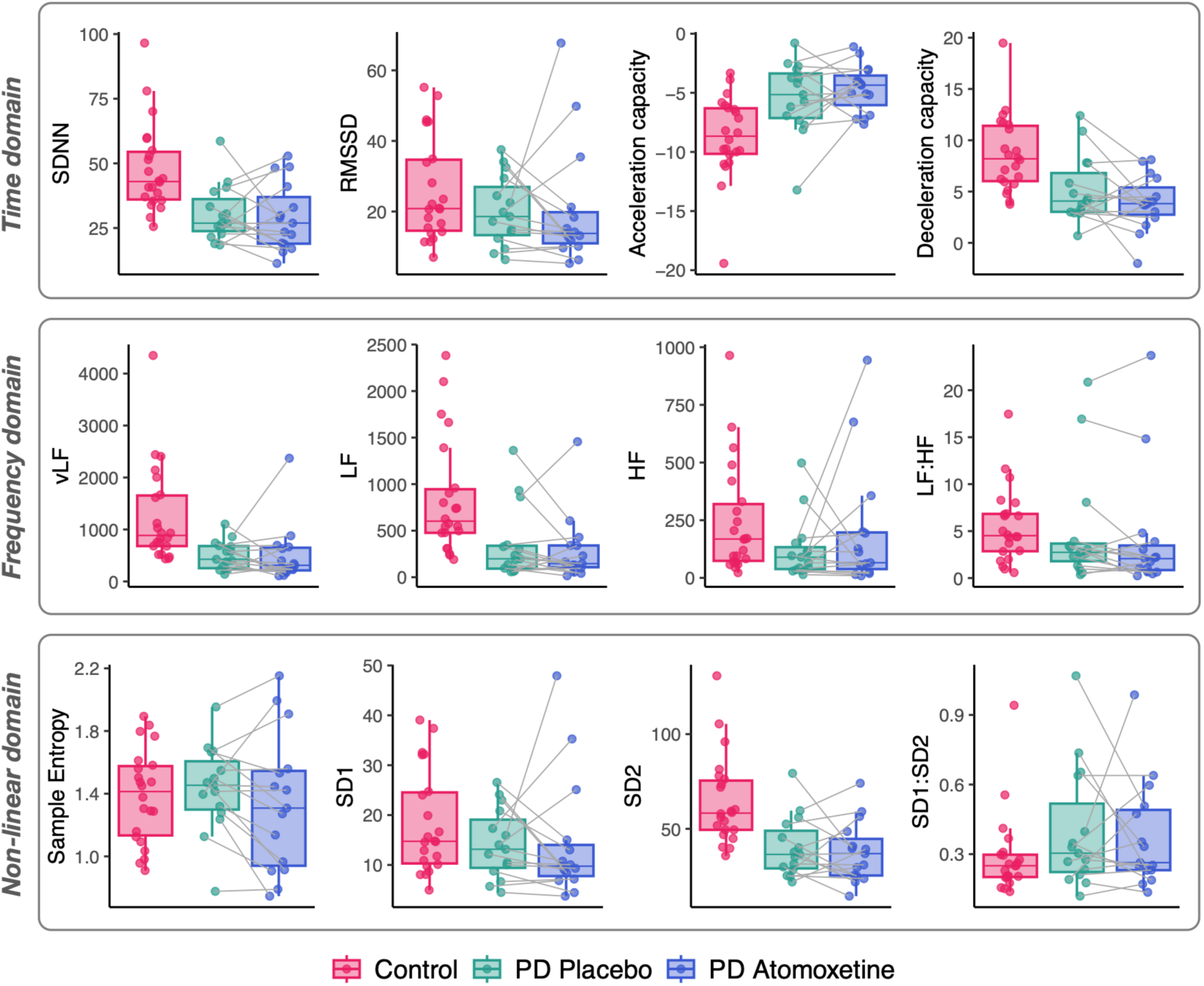
Subject-level values for HRV metrics. SDNN = Standard deviation of RR intervals, RMSSD = Square root of the mean squared differences of successive RR intervals, vLF = Power in the very low frequency range, LF = Power in the low frequency range, HF = Power in the high frequency range, LF:HF = LF to HF ratio, SD1 = Standard deviation of ellipse width on the Poincaré plot, SD2 = Standard deviation of ellipse length on the Poincaré plot, SD1:SD2 = SD1 to SD2 ratio.

**Figure 2.**
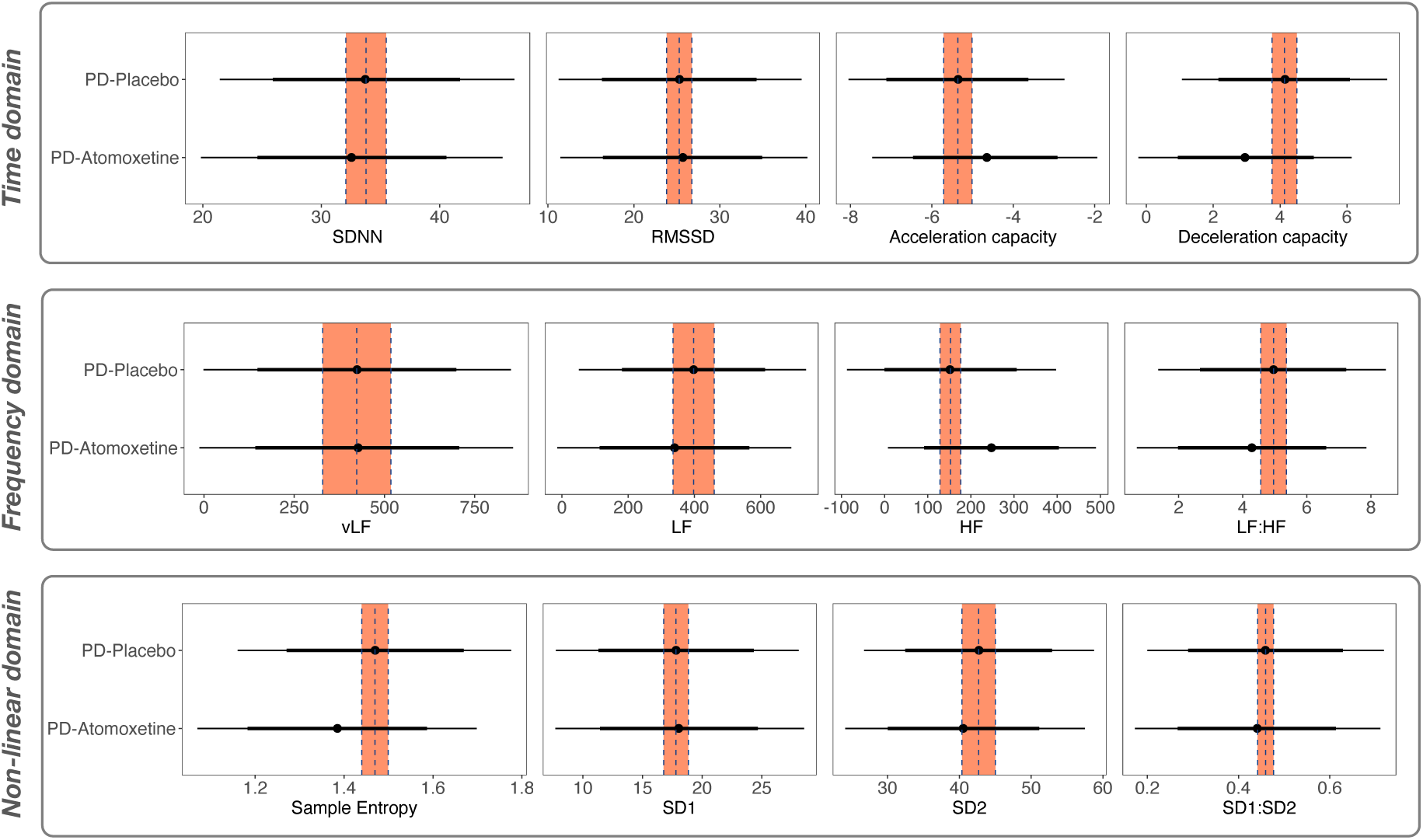
HRV comparisons on placebo vs. atomoxetine in Parkinson’s disease. Distributions (i.e., thin black lines) show the Bayesian posterior estimates of the HRV scores of patients on placebo and atomoxetine, with the Region of Practical Equivalence (ROPE) as a shaded orange column. The mean of the Placebo group is represented by the dotted vertical line. The 75% credible interval is denoted by the thick bars and the 95% credible interval by the thin bars.

In the time domain, between the atomoxetine and placebo conditions there was convincing evidence for no difference in both SDNN (effect size = −0.12, 95% CI = −8.65 to 6.00) and RMSSD (effect size = 0.03, 95% CI = −8.08 to 8.76), with the posterior means falling within the ROPE. Evidence was also in favour of no group difference for both Acceleration (effect size = 0.30, 95% CI = −0.92 to 2.26) and Deceleration (effect size = −0.46, 95% CI = −2.92 to 0.61) capacity, with the credible intervals of the two groups largely overlapping.

In the frequency domain, there was clear evidence for no difference in vLF power (effect size = 0.005, 95% CI = −249.73 to 254.08) and LF power (effect size = −0.22, 95% CI = −248.78 to 139.84), with the group means falling within the ROPE. Evidence was also indicative of a lack of group difference for HF power (effect size = 0.46, 95% CI = −38.02 to 225.46) and the LF:HF ratio (effect size = −0.39, 95% CI −0.57 to 0.19) given the considerable overlap of the credible intervals.

In the non-linear domain, there was good evidence of no difference for SD1 (effect size = 0.03, 95% CI = −5.69 to 6.21) and SD2 (effect size = −0.16, 95% CI = −11.63 to 7.31) with the means falling within the ROPE. Evidence was also suggestive of a lack of group difference for both Sample Entropy (effect size = −0.33, 95% CI = −0.25 to 0.09) and SD1:SD2 (effect size = −0.09, 95% CI = −0.17 to 0.13), with largely overlapping credible intervals.

### Evidence for altered heart rate variability in Parkinson’s disease compared to healthy controls

We found evidence of group differences in HRV scores across multiple measures in participants with Parkinson’s disease on placebo compared to healthy controls.

In the time domain, there was strong evidence of group differences in the Parkinson’s group vs. controls for SDNN (effect size = −1.03, 95% CI = −25.29 to −5.75), Acceleration Capacity (effect size = 0.87, 95% CI = 0.76 to 5.15) and Deceleration Capacity (effect size = −0.88, 95% CI = −5.46 to −0.79), with the Parkinson’s disease group mean and the 95% credible interval falling entirely outside of the ROPE and non-overlapping with the control group. In contrast, there was evidence in favour of no group difference for RMSSD (effect size = −0.40, 95% CI = −13.62 to 3.09) given the considerable overlap of the credible intervals.

In the frequency domain there was good evidence for reduced vLF power in Parkinson’s disease patients compared to controls (effect size = −0.90, 95% CI = −1194.72 to −190.02), with the mean of the posterior falling well outside the ROPE and non-overlapping credible intervals. In contrast, evidence was in favour of no group difference for LF power (effect size = −0.38, 95% CI = −580.79 to 47.77), HF power (effect size = −0.53, 95% CI = −247.41 to 22.27) and LF:HF score (effect size = −0.12, 95% CI = −3.62 to 2.48), with credible intervals falling within the ROPE and overlapping.

In the non-linear domain, we found strong evidence of reduced SD2 in the Parkinson’s disease group (effect size = −1.04, 95% CI = −35.80 to −8.51), with no overlap in the credible intervals. Whereas evidence suggested a lack of group differences in SD1 (effect size = −0.39, 95% CI = −9.52 to 2.27), SD1:SD2 (effect size = 0.41, 95% CI = −0.04 to 0.22) and Sample Entropy (effect size = −0.14, 95% CI = −0.14 to 0.23), with overlapping credible intervals that crossed into the ROPE.

**Figure 3.**
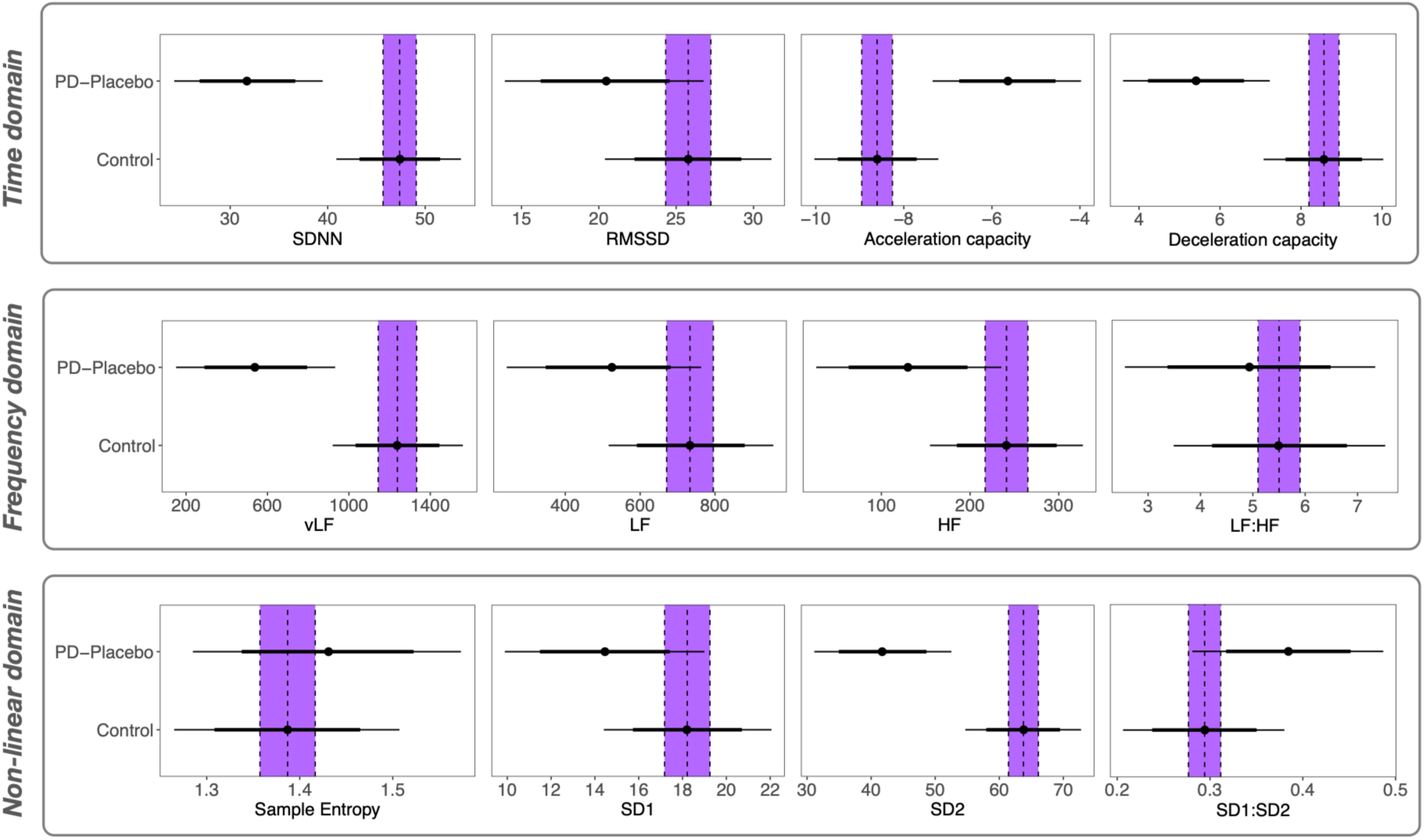
HRV comparisons between Parkinson’s disease (on placebo) and healthy controls. Distributions (i.e., thin black lines) show the Bayesian posterior estimates of the HRV scores of patients with Parkinson’s disease (on placebo) and healthy controls, with the Region of Practical Equivalence (ROPE) as a shaded purple column. The mean of the Control group is represented by the dotted vertical line. The 75% credible interval is denoted by the thick bars and the 95% credible interval by the thin bars.

### Caudal locus coeruleus integrity relates to measures of HRV in Parkinson’s disease

In the exploratory *post hoc* analysis, we tested whether there was a relationship between caudal locus coeruleus integrity and the HRV metrics that were altered in Parkinson’s disease relative to controls (i.e., SDNN, AC, DC, vLF and SD2). We found moderate evidence for a positive correlation between caudal locus coeruleus contrast-to-noise ratio (CNR) and both vLF power (R median = 0.38, BF = 2.65) and SD2 (R median = 0.41, BF = 3.08) (See Figure 4). That is, higher caudal locus coeruleus integrity related to stronger power in the vLF band and larger SD2. We did not find evidence of a relationship between caudal locus coeruleus CNR and SDNN (R median = 0.34, BF = 1.78), AC (R median = −0.34, BF = 1.76) or DC (R median = 0.21, BF = 0.822).

**Figure 4.**
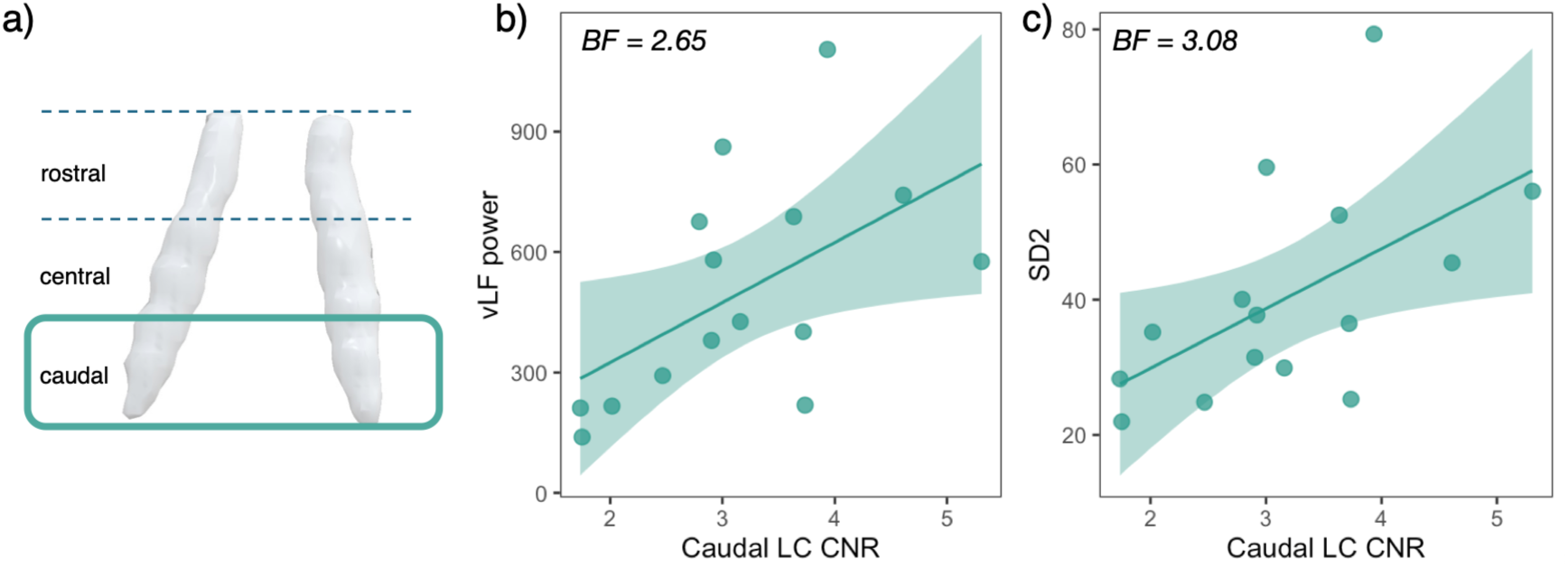
Relationship between locus coeruleus integrity and HRV. a) We extracted the contrast-to-noise ratio from the caudal portion of the locus coeruleus as a measure of integrity in Parkinson’s disease participants. b) Relationship between vLF power and caudal locus coeruleus integrity in Parkinson’s disease. c) Relationship between SD2 and caudal locus coeruleus integrity in Parkinson’s disease. CNR = contrast-to-noise ratio; LC = locus coeruleus.

## Discussion

We report evidence for the preservation of short-term heart rate variability (HRV) following 40 mg atomoxetine in people with Parkinson’s disease. Several measures of HRV were altered in Parkinson’s disease relative to healthy controls. Consistent with the role of the locus coeruleus in modulating cardiac function, reduced caudal locus coeruleus integrity was associated with abnormal HRV.

Atomoxetine did not alter short-term HRV in people with Parkinson’s disease, nor change pulse rates and sitting blood pressure. There was a small elevation of diastolic blood pressure lying down. The absence of cardiac effects is important in considering safety, made especially relevant by the evidence that the same atomoxetine dose (40 mg) is sufficient to modulate behaviour and restore brain activity and connectivity in Parkinson’s disease.^4–6,8,46^ Encouragingly, this suggests that 40 mg atomoxetine may benefit cognitive and psychiatric symptoms in Parkinson’s disease without affecting cardiac and cardiovascular regulation. The noradrenergic system has a diverse influence on cognitive processes and behavioural regulation.^47–49^ Disruptions to this system contribute to cognitive and psychiatric symptoms in neurodegenerative diseases of ageing.^2^ With growing recognition of a noradrenergic subtype in Parkinson’s disease,^1^ there is scope for repurposed noradrenergic drugs, such as atomoxetine, to form part of a personalised treatment approach in Parkinson’s disease and related conditions.^3^

Previous work has identified HRV abnormalities in Parkinson’s disease compared to healthy controls, although there is considerable variability in findings across and within studies.^23,50^ Heart rate is regulated by the sympathetic and parasympathetic autonomic nervous systems, and their interaction with cardiovascular (e.g., baroreceptors) and respiratory regulatory mechanisms.^51^ Optimal levels of heart rate variability mean an organism can adaptively respond to whatever their current context requires, with reduced HRV a feature of many health problems and diseases.^52^ We found reduced HRV in Parkinson’s disease across measures from the time- and frequency-domains (i.e., SDNN and vLF) and in a complexity measure (SD2). Measures of deceleration-related and acceleration-related capacities of heart rate were also altered compared to controls. Reductions in various HRV measures, as well as a reduced deceleration capacity, have been previously shown in Parkinson’s disease,^23,53–55^ consistent with the widespread pathological changes that impact both central and peripheral regulators of heart rate variability.^18^

The locus coeruleus plays a key role in cardiac regulation. It provides direct noradrenergic modulation over preganglionic sympathetic neurons in the intermediolateral cell column of the spinal cord and over preganglionic parasympathetic vagal neurons, including the dorsal motor nucleus of the vagus and the nucleus ambiguous.^21^ The locus coeruleus also exerts influence via projections to sympathetic premotor nuclei, including those in the rostroventrolateral medulla that contain an intrinsic pacemaker activity and are involved in maintaining blood pressure and heart rate.^56^ Focusing on the measures that were altered in people with Parkinson’s disease compared to controls, we found an association between caudal locus coeruleus contrast-to-noise ratio and certain HRV measures (i.e., very low frequency and complexity). Specifically, reduced caudal locus coeruleus integrity was associated with reduced HRV in those measures. Locus coeruleus contrast on neuromelanin-sensitive imaging has also been associated with high frequency HRV in healthy people.^57^ Taken together, neuromelanin-sensitive imaging of the locus coeruleus is emerging as a useful biomarker in psychiatry and neurology that can signal both cognitive and autonomic and changes,^58^ which may help predict the patients who will respond best to noradrenergic therapy.^7^

Our key finding is that short-term heart rate variability was preserved in people with Parkinson’s disease after an atomoxetine challenge. This favourable signal will benefit from replication in a larger sample with longer-duration ECG recordings, and chronic atomoxetine administration. Nevertheless, the results are timely given the growing interest in repurposing atomoxetine to treat cognitive and psychiatric symptoms in neurodegenerative diseases of ageing — as demonstrated by the work in Parkinson’s disease, current trials in Parkinson-plus disorders^59^ and mild cognitive impairment,^10^ and recent calls for trials in Alzheimer’s disease.^60,61^ Our findings are in favour of a safe cardiovascular profile for atomoxetine in Parkinson’s disease, lending further support to noradrenergic modulation as a viable treatment strategy for neuropsychiatric symptoms in Parkinson’s disease and related disorders.

## Data availability

Code and data to reproduce manuscript figures and statistical analyses are openly available [note: link to be made public on publication].

## Acknowledgements

We thank all volunteers and their families for their participation, all staff at the Wolfson Brain Imaging Centre and NIHR Cambridge Clinical Research Facility for their help with data collection.

