## Supplementary material for "Short term heart rate variability is preserved in Parkinson’s disease under atomoxetine"


1. Heart rate variability metrics Page 2

2. Locus coeruleus imaging Page 4

3. Within session physiological effects Page 7

4. Within session subjective effects Page 9

5. Group mean and standard deviation of HRV measures Page 11

6. References Page 12

### 1. Heart rate variability metrics

We computed heart rate variability measures under the time and frequency domain, as well as calculating nonlinear complexity measures, using the PhysioNet Cardiovascular Signal Toolbox [1].

In the *time domain*, we obtained the standard deviation of normal-to-normal (NN) intervals (SDNN), and the square root of the mean of the sum of the squares of differences between successive NN intervals (RMSSD) indicating beat-to-beat variance in heart rate. Acceleration (AC) and deceleration (DC) capacities were also quantified with phase-rectified signal averaging.

In the *frequency domain* spectral analyses were performed to measure relative power, i.e., milliseconds squared divided by cycles per second ($ms^2$/Hz) then divided by the summed absolute power of each frequency band. The toolbox employs Lomb periodogram transformations to estimate relative power and separates signal into their according band. We derived power in the high frequency (HF) range (0.15-0.4 Hz), low frequency (LF) range (0.04-0.15 Hz) and very low frequency (vLF) range (0.003-0.04 Hz). We also calculated the LF:HF ratio.

In the *non-linear domain* sample entropy (SampEn) was calculated as a measure of nonlinear complexity, with higher sample entropy indicating less regularity and more complexity in the time-series. A Poincaré plot (i.e., a map of every RR interval against the prior interval) was also captured and analysed by fitting an ellipse to derive other nonlinear measurements of heart rate variability. SD1 is the standard deviation of the distance of each point from the $y = x$ axis (ellipse width) and captures short-term heart rate variability. SD2 is the standard deviation of the distance of each point from the $y = x + average\ RR$ (ellipse length) and captures both short- and long-term heart rate variability. From this we computed the ratio SD1:SD2.

***PhysioNet cardiovascular signal toolbox parameter settings***

We analysed the 10-minute ECG recording as a single block to enhance feature extraction. '*HRVparams.RejectionThreshold*' was increased from the default 20% to 35%, meaning that 35% of data within the window can be rejected before it is considered too low quality. The maximum percentage of missing data, '*HRVparams.MissingDataThreshold*', was increased from the default 15% to 35%. The signal quality index (SQI) ‘*HRVparams.sqi.LowQualityThreshold*’ was also lowered from 90% to 60%. SQI is calculated using a moving window of 10-seconds across the entire signal.

Raw signals were converted to RR intervals with the ‘*ConvertRawDataToRRIntervals.m*’ function. Atrial fibrillation and premature ventricular contraction (PVC) detection and removal were carried out using the automated ‘*HRVparams.af.on*’ and ‘*HRVparams.PVC.qrsth*’ functions respectively. Once filtering of low-quality data was completed, we interpolated the signal to complete the RR interval series with the linear default resampling.

### 2. Locus coeruleus imaging

MRI acquisition and preprocessing, and locus coeruleus signal extraction, were conducted based on pipelines reported in [2,3].

***MRI acquisition***

All patients and controls underwent MR imaging. Two controls were excluded from further imaging analysis due to incidental structural abnormalities. MR images were acquired with a 7T Magnetom Terra scanner (Siemens, Erlangen, Germany), using a 32-channel receive and circularly polarised single-channel transmit head coil (Nova Medical, Wilmington, USA). We used a 3-D high-resolution magnetisation transfer-weighted turbo flash (MT-TFL) sequence for imaging the locus coeruleus (based on [4]). 112 axial slices were used to cover both the midbrain and the pontine regions. The sequence applied a train of 20 Gaussian-shape RF-pulses at 6.72 ppm off resonance, 420º flip-angle, followed by a turbo-flash readout (TE = 4.08 ms, TR = 1251 ms, flip-angle = 8°, voxel size = 0.4 x 0.4 x 0.5 $mm^3$, 6/8 phase and slice partial Fourier, bandwidth = 140 Hz/px, no acceleration, 14.3%-oversampling, TA ~ 7 min). For each subject, the transmit voltage was adjusted based on the average flip angle in the central area of the pons obtained from a B1 pre-calibration scan. The MT-TFL sequence was repeated twice and averaged offline to improve signal-to-noise ratio. An additional scan (MT-off) was acquired with the same parameters as above but without the off-resonance pulses. A high resolution isotropic T1-weighted (T1-w) MP2RAGE image was also acquired sagittally for anatomical coregistration using the UK7T Network harmonised protocol [5]: TE = 2.58 ms, TR = 3500 ms, BW = 300 Hz/px, voxel size = 0.7 x 0.7 x 0.7 $mm^3$, FoV = 224 x 224 x 157 $mm^3$, acceleration factor (A>>P) = 3, flip angles = 5/2° and inversion times (TI) = 725/2150 ms for the first/second images.

***Image processing and coregistration pipeline***

Image processing and coregistration was based on the pipeline described in [3]. The Advanced Normalization Tools (ANTs v2.2.0) software and in-house MATLAB scripts were used for image pre-processing and the standardisation of MT images. MT images were first N4 bias field corrected for spatial inhomogeneity (number of iterations at each resolution level: 50x50x30x20, convergence threshold: $1x10^{-6}$, isotropic sizing for b-spline fitting: 200 [6] then averaged using the customised antsMultivariateTemplateConstruction2 function for improvements in signal-to-noise ratio. The T1-w MP2RAGE data were generated offline from the complex images [7]. T1-w skull-stripped images were obtained after tissue type segmentation and reconstruction using SPM12 (v7219) (http://www.fil.ion.ucl.ac.uk/spm/software/spm12/).

The pre-processed MT-weighted and T1-w images were then entered into a T1-driven, cross modality coregistration pipeline to warp the individual MT and MT-off images to the isotropic 0.5 mm ICBM152 (International Consortium for Brain Mapping) T1-w asymmetric template [8]. The individual T1-w images were first coregistered to the MT image with rigid only transformation. The MT-off image was used as the intermediate step for bridging the two modalities because the MT-off image shares similar tissue-specific contrasts with both T1-w MP2RAGE and MT-on images.

In parallel, an unbiased study-wise T1-w structural template was created using individual skull-stripped T1-w images from all controls and patients. Native T1-w images were rigid and affine transformed, followed by a hierarchical nonlinear diffeomorphic step at five levels of resolution, repeated by six runs to improve convergence. Max iterations for each resolution from the coarsest level to the full resolution were 100x100x70x50x20 (shrink factors: 10x6x4x2x1, smoothing factors: 5x3x2x1x0 voxels, gradient step size: 0.1 mm). The Greedy Symmetric Normalisation (SyN) was adopted for the transformation model of the deformation step [9]. The resulting T1-w group template was then registered to the standard ICBM152 T1-w brain following the similar rigid-affine-SyN steps at four resolution levels (max iterations: 100x70x50x50, convergence threshold: $1x10^{-6}$, shrink factors: 8x4x2x1, smoothing factors: 3x2x1x0 voxels). For all the above registration steps, cross-correlation was used for similarity metrics estimation as it performs better for linear and non-linear components during intra-modality registration. Four steps of deformations were estimated as follows (in order): MT-off to MT, T1-w to MT-off, T1-w to T1-w group template and T1-w group template to ICBM152 T1-w template. These parameters were then used as the roadmap for MT image standardisation to the ICBM brain in one step. A trilinear interpolation method was selected to preserve the absolute location and relative contrast of the signal.

***Independent probabilistic locus coeruleus atlas creation***

To facilitate accurate extraction of the locus coeruleus signal we created a study-specific unbiased locus coeruleus atlas. To do this, we used an independent sample of 29 age- and education matched healthy controls (13 female; age mean (SD) = 67 (8.2), age range = 52-84) collected under the same neuroimaging protocol. We used a validated pipeline for locus coeruleus atlas construction described in [3]. Briefly, for each axial slice on the rostrocaudal extent, the locations of the left and right locus coeruleus were determined using a semi-automated segmentation method. A threshold was defined as five standard deviations above the mean intensity in the central pontine reference region. After applying the threshold, locus coeruleus voxels on axial planes were automatically segmented into binarised images and then averaged to construct a probabilistic atlas. The independent locus coeruleus atlas generated for this study had very high similarity in the spatial distribution of probabilities and contours relative to the validated 7T locus coeruleus atlas in [3].

***Locus coeruleus signal extraction***

As a measure of locus coeruleus integrity, we quantified contrast by establishing the CNR with respect to a reference region in the central pons. A CNR map was computed voxel-by-voxel on the average MT image for each subject using the signal difference between a given voxel (V) and the mean intensity in the reference region ($Mean_{REF}$) divided by the standard deviation ($SD_{REF}$) of the reference signals ($CNR = \frac{V - Mean_{REF}}{SD_{REF}}$). CNR values were extracted bilaterally on the CNR map by applying the independent locus coeruleus probabilistic atlas (5% probability version). We computed mean CNR values for the rostral, middle and caudal portions of the left and right locus coeruleus. Comparisons of locus coeruleus CNR between the Parkinson's disease and control groups are reported in detail in O'Callaghan et al. [2].

### 3. Within session physiological effects

As described in detail below, there was no evidence of increased pulse rates under atomoxetine in either the upright or supine positions. Systolic blood pressure (upright and supine) did not increase under atomoxetine. There was a slight increase in supine diastolic blood pressure under atomoxetine, which was not observed in the upright position. There was evidence for an effect of time on upright diastolic blood pressure, which was slightly elevated at completion of testing relative to arrival and two-hours post tablet administration (for both atomoxetine and placebo). Mean values, standard deviations and ranges for blood pressure and pulse rates are shown in Supplementary Table 1.

*Pulse rates*

Both upright and supine (lying down) pulse rates did not change significantly under atomoxetine vs. placebo, as evidenced by a lack of main effect (Upright: ($F_{(1, 13.14)} = 0.50$, $p = 0.491$; BF = 1.015); Supine: $F_{(1, 13.42)} = 0.02$, $p = 0.900$; BF = 0.452), and did not vary across the three time points (i.e., arrival, two-hours post tablet, on completion of testing; Upright: $F_{(2, 63.92)} = 0.24$, $p = 0.789$; BF = 0.102); Supine: $F_{(2, 65.00)} = 0.31$, $p = 0.731$; BF = 0.172)). There was no significant interaction between drug status and time point on pulse rate (Upright: $F_{(2, 63.92)} = 0.49$, $p = 0.616$; BF = 0.063; Supine: $F_{(2, 65.00)} = 0.30$, $p = 0.741$; BF = 0.023).

*Blood pressure*

We did not find a significant main effect of atomoxetine on upright systolic and diastolic blood pressure (systolic: $F_{(1, 13.35)} = 0.48$, $p = 0.499$; BF = 0.250; diastolic: $F_{(1, 13.67)} = 2.15$, $p = 0.165$; BF = 0.262). There was no main effect of timepoint on upright systolic blood pressure (systolic: $F_{(2, 63.95)} = 2.77$, $p = 0.070$; BF = 1.750), however there was an effect on upright diastolic blood pressure ($F_{(2, 64.09)} = 4.36$, $p = 0.017$; BF = 3.034). This effect was driven by increased diastolic blood pressure on completion of testing, compared to arrival and two hours post testing (arrival vs. completion: t(65) = 2.442, p = 0.026, BF = 0.546); two hours post vs. completion: t(65) = 3.741, p = 0.001, BF = 1.274)). There was no significant interaction effects between condition and timepoint on upright blood pressure (systolic: $F_{(2, 63.95)} = 0.21$, $p = 0.813$; BF = 0.111; diastolic: $F_{(2, 64.09)} = 0.51$, $p = 0.602$; BF = 0.170).

We did not find a significant effect of atomoxetine on supine (lying down) systolic blood pressure ($F_{(1, 13.35)} = 0.53$, $p = 0.481$; BF = 1.711), however we found some evidence of a higher diastolic blood pressure on atomoxetine in the supine position ($F_{(1, 13.47)} = 0.10$, $p = 0.757$; BF = 9.664). There was no significant main effect of timepoint on supine blood pressure (systolic: $F_{(2, 65.00)} = 0.92$, $p = 0.405$; BF = 0.523; diastolic: $F_{(2, 65.00)} = 2.86$, p = 0.065; BF = 0.494). We found no interaction effect of condition and time point on supine blood pressure (systolic: $F_{(2, 65.00)} = 0.18$, $p = 0.833$; BF = 0.179; diastolic: $F_{(2, 65.00)} = 0.36$, $p = 0.702$; BF = 1.051).

*Supplementary Table 1*

| **Measure** | | | **Placebo** | **Atomoxetine** |
|---|---|---|---|---|
| Pulse rates | Lying | Arrival | 69.86 (48 – 83; 9.16) | 69.27 (56 – 90; 9.77) |
| | | 2-hours | 68.93 (55 – 84; 10.39) | 74.60 (58 – 95; 11.33) |
| | | Completion | 67.27 (50 – 85; 8.43) | 70.33 (50 – 93; 11.39) |
| | Upright | Arrival | 75.20 (49 – 100; 13.74) | 72.93 (56 – 110; 14.27) |
| | | 2-hours | 70.26 (54 – 86; 9.29) | 78.80 (60 – 106; 14.51) |
| | | Completion | 68.86 (49 – 80; 8.51) | 78.40 (57 – 116; 16.49) |
| Systolic blood pressure | Lying | Arrival | 129.27 (84 – 151; 17.86) | 133.73 (113 – 175; 17.61) |
| | | 2-hours | 127.07 (112 – 156; 13.54) | 135.33 (114 – 169; 16.16) |
| | | Completion | 134.00 (116 – 155; 12.59) | 144.00 (111 – 186; 19.93) |
| | Upright | Arrival | 127.73 (80 – 165; 19.88) | 130.33 (102 – 166; 19.15) |
| | | 2-hours | 124.20 (114 – 145; 10.24) | 130.87 (93 – 167; 19.93) |
| | | Completion | 141.29 (118 – 176; 15.26) | 137.60 (97 – 183; 23.13) |
| Diastolic blood pressure | Lying | Arrival | 70.40 (50 – 84; 9.50) | 74.07 (55 – 85; 7.42) |
| | | 2-hours | 67.60 (58 – 80; 5.08) | 74.33 (58 – 94; 8.50) |
| | | Completion | 72.67 (59 – 87; 8.00) | 78.00 (63 – 98; 10.43) |
| | Upright | Arrival | 72.00 (43 – 82; 10.70) | 73.27 (56 – 83; 7.68) |
| | | 2-hours | 68.53 (51 – 81; 8.62) | 71.80 (52 – 93; 10.39) |
| | | Completion | 77.64 (61 – 88; 8.10) | 76.47 (61 – 88; 8.74) |

Mean values, standard deviations and ranges for blood pressure and pulse rates on Atomoxetine and Placebo. Data are presented as mean (range; SD).

### 4. Within session subjective effects

Although the visual analogue scale (VAS) is a continuous measure, participants often respond at either end of the scale, leading to bi- or even tri-modal distributions (Supplementary Figure 1). Such dynamics are not well captured by conventional analyses (e.g., linear regression) that assume multivariate normality. To address this issue, we analysed the VAS data using a Bayesian ordered beta regression model [10]. The strength of this model is that it simultaneously estimates the probability of responses at the scale's lower and upper bounds as well as continuously distributed responses in between the bounds. As described below, there was no change in subjective mood/arousal levels within the sessions.

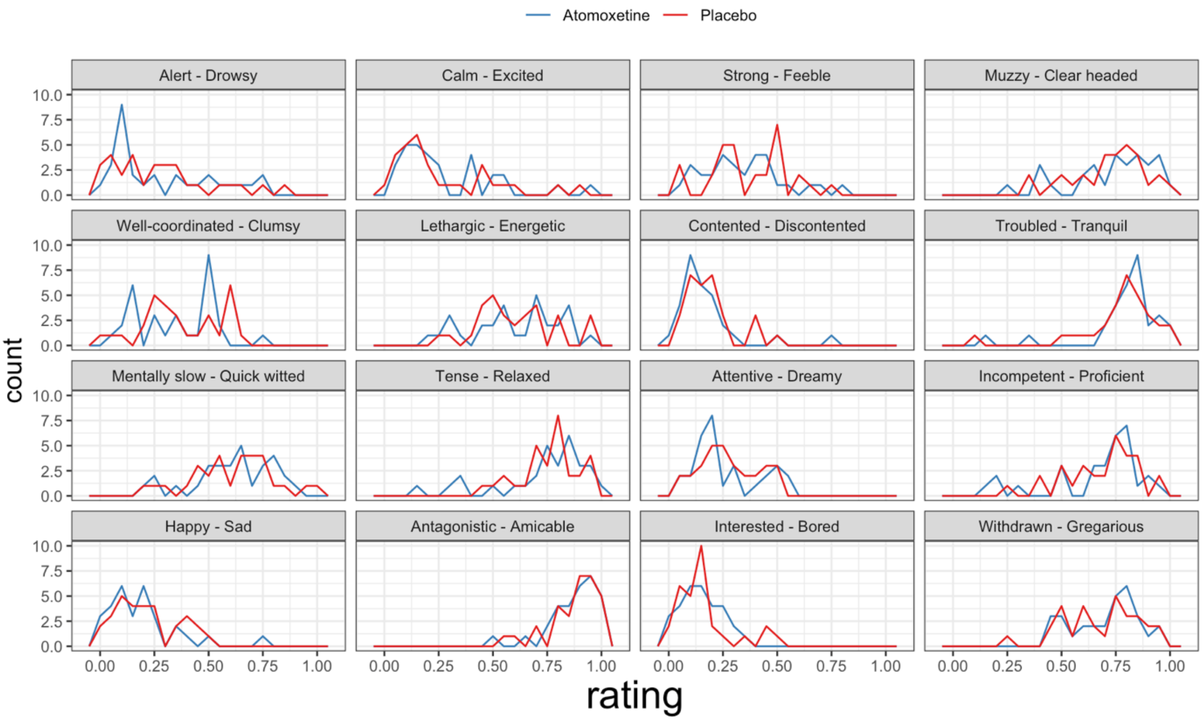


**Supplementary Figure 1.** Frequency polygons of VAS ratings in the PD group. Each panel represents one VAS item, as described by the panel titles. The first term of each panel title corresponds to the left extreme of that VAS item (i.e. rating = 0), whereas the second term corresponds to the right extreme (i.e. rating = 1).

We modelled drug (atomoxetine vs. placebo), time point (2 hours post administration vs. baseline), the drug × time interaction, and session (first vs. second visit) as categorical predictors of the VAS response (i.e., fixed effects), and we allowed the intercept to vary by VAS item and by participant (i.e., random effects). We assigned a weakly informative normal prior on the regression coefficients $\beta \sim N(0, 5)$ [10]. For posterior inference, we set a region of practical equivalence (ROPE) at $\pm 0.1 \times SD_{VAS} = \pm 0.019$, corresponding to a negligible effect size [11,12].

There were no main effects of drug or time point on VAS response, as the posterior distributions of these coefficients were largely contained by the ROPE (Supplementary Figure 2; drug: β = -0.01, 95% HDI [-0.06, 0.03], proportion in ROPE = 52.39%; time point: β = 0.04, 95% HDI [-0.01, 0.08], proportion in ROPE = 20.69%). Although the posterior estimate of the drug × time point interaction effect was greater than the upper bound of the ROPE, we failed to reject the null as a relatively large proportion of the posterior distribution was contained by the ROPE (interaction: β = 0.03, 95% HDI [-0.01, 0.08], proportion in ROPE = 27.63%). Taken together, these results suggest that atomoxetine did not induce a significant change in subjective states, as measured by the VAS.

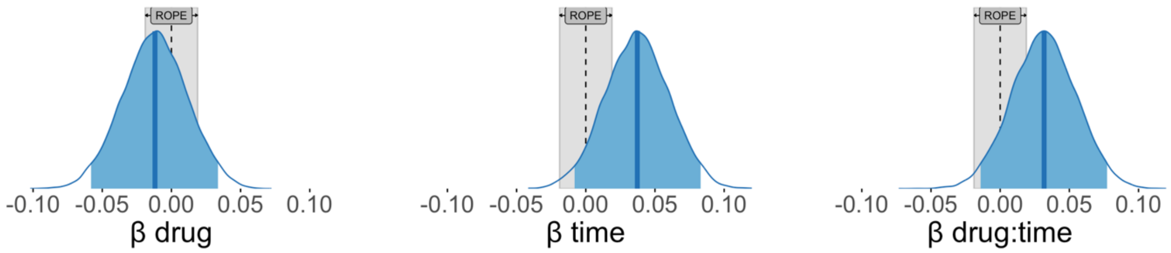


**Supplementary Figure 2.** Posterior distributions of predictors of VAS responses. For each panel, the dark blue vertical line represents the median – that is, the posterior estimate of the regression coefficient; the blue shaded area represents the 95% highest density interval of the posterior distribution; and the blue density trace represents the full posterior distribution. The grey area represents a region of practical equivalence (ROPE), corresponding to a negligible effect size of ±0.1.

### 5. Group mean and standard deviation of HRV measures

*Supplementary Table 2*

| | | Control | PD-Placebo | PD-Atomoxetine |
|---|---|---|---|---|
| Time Domain | SDNN | 47.74 (17.02) | 30.70 (10.79) | 29.15 (13.09) |
| | RMSSD | 25.91 (14.63) | 20.09 (9.80) | 20.33 (17.45) |
| | Acceleration Capacity | -8.69 (3.49) | -5.41 (3.05) | -4.66 (2.00) |
| | Deceleration Capacity | 8.67 (3.70) | 5.17 (3.23) | 3.93 (2.68) |
| Frequency Domain | Very Low | 1252.93 (945.30) | 500.85 (277.57) | 505.64 (567.87) |
| | Low | 840.09 (624.32) | 351.25 (388.96) | 286.69 (362.10) |
| | High | 244.49 (240.35) | 122.08 (132.41) | 193.99 (271.33) |
| | Low: High | 5.54 (4.01) | 4.85 (6.03) | 4.17 (6.48) |
| Non-linear | SD1 | 18.34 (10.36) | 14.22 (6.94) | 14.39 (12.37) |
| | SD2 | 64.44 (23.33) | 40.28 (15.80) | 37.74 (16.19) |
| | SD1:SD2 | 0.29 (0.17) | 0.40 (0.26) | 0.37 (0.23) |
| | Sample Entropy | 1.39 (0.30) | 1.43 (0.28) | 1.34 (0.44) |

Mean and standard deviations for HRV measures in healthy controls, Parkinson’s disease patients on Placebo and Atomoxetine. Data are presented as mean (SD).
